# NON-WHITE ETHNICITY, MALE SEX, AND HIGHER BODY MASS INDEX, BUT NOT MEDICATIONS ACTING ON THE RENIN-ANGIOTENSIN SYSTEM ARE ASSOCIATED WITH CORONAVIRUS DISEASE 2019 (COVID-19) HOSPITALISATION: REVIEW OF THE FIRST 669 CASES FROM THE UK BIOBANK

**DOI:** 10.1101/2020.05.10.20096925

**Authors:** Zahra Raisi-Estabragh, Celeste McCracken, Maddalena Ardissino, Mae S. Bethell, Jackie Cooper, Cyrus Cooper, Nicholas C. Harvey, Steffen E. Petersen

**Author notes:** **Correspondence:** Professor Steffen E. Petersen. William Harvey Research Institute, NIHR Barts Biomedical Research Centre, Queen Mary University of London, London, UK.

## Abstract

**Background:** Cardiometabolic morbidity and medications, specifically Angiotensin Converting Enzyme inhibitors (ACEi) and Angiotensin Receptor Blockers (ARBs), have been linked with adverse outcomes from coronavirus disease 2019 (COVID-19). This study aims to investigate factors associated with COVID-19 positivity for the first 669 UK Biobank participants; compared with individuals who tested negative, and with the untested, presumed negative, rest of the population.

**Methods:** We studied 1,474 participants from the UK Biobank who had been tested for COVID-19. Given UK testing policy, this implies a hospital setting, suggesting at least moderate to severe symptoms. We considered the following exposures: age, sex, ethnicity, body mass index (BMI), diabetes, hypertension, hypercholesterolaemia, ACEi/ARB use, prior myocardial infarction (MI), and smoking. We undertook comparisons between: 1) COVID-19 positive and COVID-19 tested negative participants; and 2) COVID-19 tested positive and the remaining participants (tested negative plus untested, n=501,837). Logistic regression models were used to investigate univariate and mutually adjusted associations.

**Results:** Among participants tested for COVID-19, non-white ethnicity, male sex, and greater BMI were independently associated with COVID-19 positive result. Non-white ethnicity, male sex, greater BMI, diabetes, hypertension, prior MI, and smoking were independently associated with COVID-19 positivity compared to the remining cohort (test negatives plus untested). However, similar associations were observed when comparing those who tested negative for COVID-19 with the untested cohort; suggesting that these factors associate with general hospitalisation rather than specifically with COVID-19.

**Conclusions:** Among participants tested for COVID-19 with presumed moderate to severe symptoms in a hospital setting, non-white ethnicity, male sex, and higher BMI are associated with a positive result. Other cardiometabolic morbidities confer increased risk of hospitalisation, without specificity for COVID-19. Notably, ACE/ARB use did not associate with COVID-19 status.

## INTRODUCTION

Coronavirus disease 2019 (COVID-19), the clinical illness caused by the severe acute respiratory syndrome coronavirus 2 (SARS-CoV-2), has reached pandemic levels^1^. There has been growing recognition that patients with underlying cardiometabolic morbidities may be suffering higher rates of infection and a more severe disease course than the general population^2–4^. Debate has ensued regarding whether these observations relate to the conditions themselves or the medications with which they are treated. In particular, some have suggested a mechanistic role for Angiotensin Converting Enzyme inhibitors (ACEi) or Angiotensin Receptor Blockers (ARBs)^5^. However, recent reports have not produced convincing evidence for the specific association of ACEi/ARBs with poorer outcomes^5-7^. Cardiometabolic diseases are common and ACEi/ARBs are used by many vulnerable patients. It is therefore important to better understand the augmented risk associated with cardiometabolic factors and ACEi/ARB use with COVID-19, to inform clinical practice, and guidance to patients.

The UK Biobank (UKB) is a large cohort study comprising data from 500,000 participants from across the UK, characterised in detail at baseline (2006-2010), and with linkages to Hospital Episode Statistic (HES) data enabling prospective tracking of health outcomes. In response to the COVID-19 pandemic, the UKB facilitated rapid release of data on COVID-19 testing for its participants, providing a unique opportunity to study the effects of many well-defined exposures on COVID-19 status. Given the contemporary UK government policy that limits COVID-19 testing to those presenting to hospital with symptoms suspicious of COVID-19, this is presumed to imply symptomatology severe enough to warrant admission to, and testing in, hospital.

The aim of this study is to examine the first-released data from the UKB to establish the association between COVID-19 positivity and demographic factors (age, sex, ethnicity), cardiometabolic factors [body mass index (BMI), diabetes, hypertension, hypercholesterolaemia, prior myocardial infarction (MI), smoking], and ACEi/ARB use.

## METHODS

### Setting and study population

UKB is a nation-wide prospective cohort study including >500,000 participants from across the UK. Individuals aged 40-69 years old identified via National Health Service (NHS) registers were recruited over a four-year period between 2006-2010. Participants underwent detailed baseline assessment including characterisation of socio-demographics, lifestyle, medical history, and a series of physical measures. The protocol is publicly available^8^. Linkages with HES data permit longitudinal tracking of health outcomes for all participants with conditions recorded according to international classification of disease (ICD) codes. In addition, UKB has produced algorithmically defined outcome data for incidence of key illness, such as MI, through integration of data from multiple sources^9^. The latest update of the COVID-19 data includes test results from 16/03/2020 to 14/04/2020. During this time period, the UK had moved away from community testing and was almost exclusively testing for COVID-19 in hospital settings. Thus, the UKB COVID-19 cohort in this time period comprises participants who were tested for SARS-CoV-2 whilst admitted to hospital, and therefore are likely to have a relatively severe presentation.

### Ethics

This study was covered by the ethics approval for UKB studies from the NHS National Research Ethics Service on 17th June 2011 (Ref 11/NW/0382) and extended on 10th May 2016 (Ref 16/NW/0274).

### Statistical analysis

Statistical analysis was performed using R Version 3.6.2 [R Core Team (2019). R: A language and environment for statistical computing. R Foundation for Statistical Computing, Vienna, Austria. URL https://www.R-project.org/], and RStudio Version 1.2.5019 [RStudio Team (2015). RStudio: Integrated Development for R. RStudio, Inc., Boston, MA URL http://www.rstudio.com/]. We considered the following exposures: age, sex, ethnicity, body mass index (BMI), diabetes, hypertension, high cholesterol, ACEi/ARB use, prevalent MI, and smoking. The cardiometabolic and demographic factors were selected based on existing reports of their potential association with COVID-19 outcomes^4,10,11^. ACEi/ARBs were considered due to reports of potential mechanistic role of these medications in the clinical course of COVID-19^5^. We used age, sex, and ethnicity (white Caucasian vs non-white Caucasian) as recorded at baseline. BMI was calculated from height and weight recorded at baseline. Smoking status was based on self-report. Hypertension, diabetes, and hypercholesterolaemia were defined through cross-checking across self-report and HES data. A list of ICD codes used is available in Supplementary Table 1. Information on prior MI was obtained from the UKB algorithmically defined health outcomes. ACEi/ARB use was determined from self-report (Supplementary Table 2). We created three cohorts: positives (test positive), confirmed negatives (test negative), and the remaining untested cohort (Figure 1). We firstly compared the COVID-19 positive cohort with the combined cohort of COVID-19 test negative and the untested UKB population. In order to investigate possible bias relating to hospitalisation status, we also considered the importance of these exposure variables in two further comparisons: test positives vs test negatives and test negatives vs untested population. We used logistic regression models to elucidate univariate and then multivariate associations. There was no evidence of multicollinearity with inflation factor (VIF) <2.0 for all covariates. We present odds ratio (OR) for each exposure with the corresponding 95% confidence interval (CI) and p-value.

**Figure 1.**
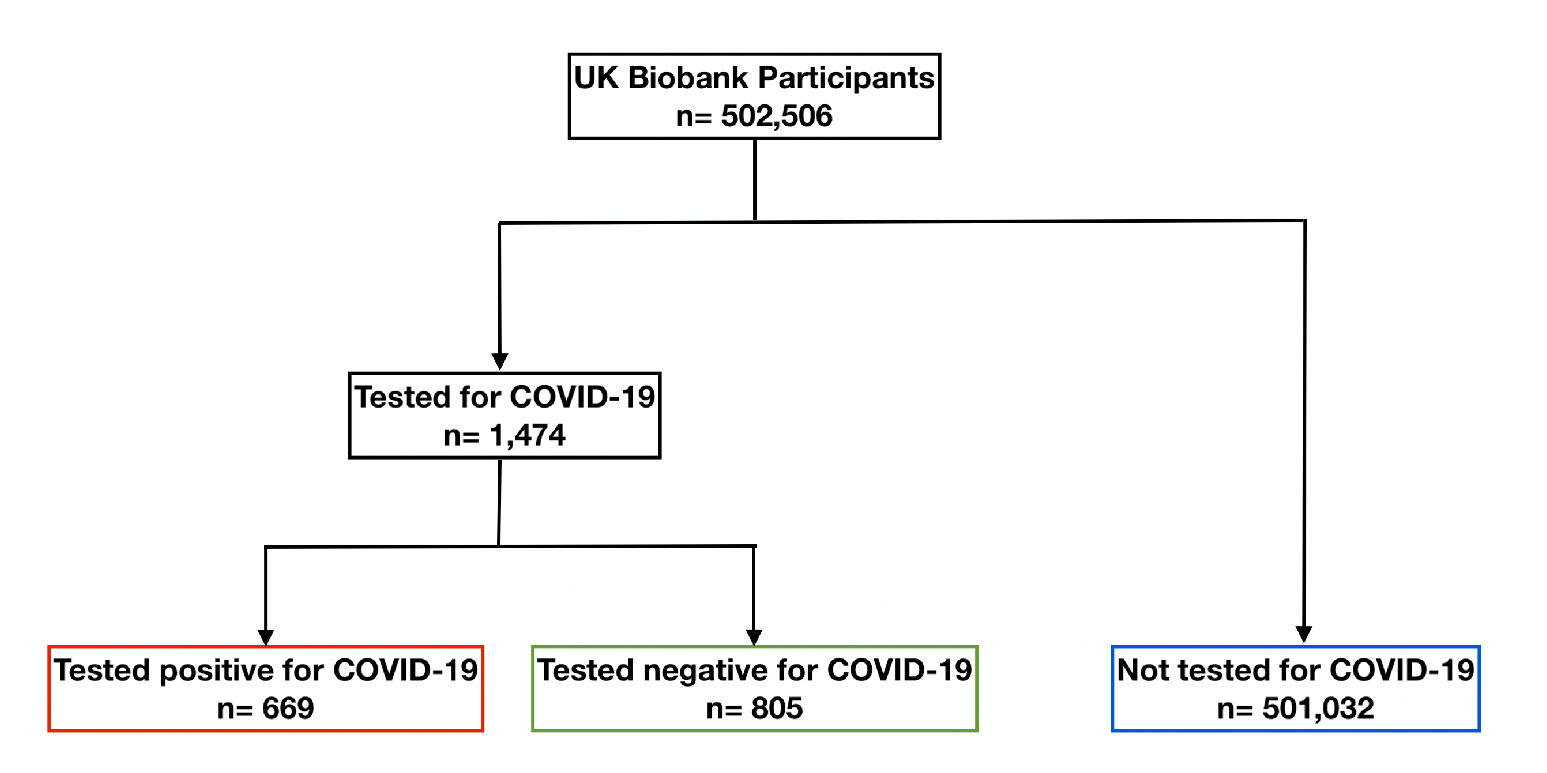
Summary of COVID-19 testing and results for UK Biobank participants. **Figure 1 legend:** Data includes COVID-19 test results from 16/03/2020 to 14/04/2020. During this time period, 1,474 participants, of the whole UK Biobank cohort (n=502,506) have been tested for COVID-19. Of those tested, 669 participants had a positive result and 805 tested negative. The remaining participants (n=501,032) have not been tested.

## RESULTS

### Baseline characteristics

The 1,474 UKB participants that had been tested for COVID-19 in the first release of UKB COVID-19 data are included in this analysis (Table 1). Among those who were tested, 669 tested positive and 805 tested negative. There was no record of testing for the remainder of the UKB cohort (n=501,032); this untested population was therefore presumed negative (Figure 1).

**Table 1.** Participant characteristics. **Table 1 caption:** Data are n (%), mean (standard deviation), or median [interquartile range]. COVID-19 data includes test results from 16/03/2020 to 14/04/2020 from hospital settings. *We report age of participants as of 01/04/2020. **smoking includes current and previous smoking. ^†^ACEi/ARB use is defined as a binary measure, defined as true if record of any of medications in supplementary Table 2. ACEi: Angiotensin Converting Enzyme inhibitor; ARB: Angiotensin Receptor Blocker; BMI: body mass index; COVID-19: coronavirus 2019.

|  | COVID-19 tested<br>(n=1,474) | COVID-19 positive<br>(n= 669) | COVID-19 negative<br>(n= 805) | Untested population<br>(n= 501,032) |
| --- | --- | --- | --- | --- |
| Sex (Male) | 787 (53.4%) | 378 (56.5%) | 409 (50.8%) | 228,335 (45.6%) |
| Age* | 69.3 (± 8.7) | 69.0 (± 8.7) | 69.5 (± 8.7) | 68.3 (± 8.1) |
| White ethnicity | 1,306 (88.6%) | 565 (84.5%) | 741 (92.0%) | 471,391 (94.1%) |
| BMI (kg/m<sup>2</sup>) | 27.8 [± 6.4] | 28.2 [± 6.3] | 27.4 [± 6.3] | 26.7 [± 5.8] |
| Smoking** | 802 (54.4%) | 356 (53.2%) | 446 (55.4%) | 224,593 (44.8%) |
| Prevalent MI | 138 (9.4%) | 56 (8.4%) | 82 (10.2%) | 20,677 (4.1%) |
| Diabetes | 229 (15.5%) | 117 (17.5%) | 112 (13.9%) | 38,909 (7.8%) |
| Hypertension | 728 (49.4%) | 336 (50.2%) | 392 (48.7%) | 174,266 (34.8%) |
| High cholesterol | 515 (34.9%) | 240 (35.9%) | 275 (34.2%) | 117,181 (23.4%) |
| ACEi/ARB use† | 312 (21.2%) | 146 (21.8%) | 166 (20.6%) | 71,284 (14.2%) |

In comparison to the untested cohort, the COVID-19 positive cohort were older, predominantly male (56.5%), and had a greater proportion of individuals with non-white ethnicities (14.6% vs 5.4%). They had an all-round poorer cardiometabolic profile, with higher BMI, and higher rates of smoking, prior MI, diabetes, hypertension and high cholesterol. They also reported greater use of ACEi/ARB agents (21.8% vs 14.2%). However, comparing the COVID-19 positive cohort with the tested negative cohort (n=805), the differences were much less pronounced, for the tested negative cohort also had a greater proportion of older individuals with a globally poorer cardiometabolic profile than the untested cohort.

### Association of exposures with COVID status

#### COVID-19 positive vs not COVID-19 positive (tested negative cohort <u>plus</u> untested cohort)

We first tested whether there were univariate associations between exposures and COVID-19 positives (n=669) vs COVID-19 negatives (including both the tested negative cohort and the untested cohort, n=501,837). Univariate associations were significant for all covariates considered. In multivariate models, the independent predictors of COVID-19 positivity were older age, male sex, non-white ethnicity, greater BMI, diabetes, hypertension, prior MI, and smoking (Table 2, Figure 2).

**Table 2.** Odds Ratios, 95% confidence intervals, and p-values for each exposure from univariate and multivariate logistic regression models in the three defined comparisons^**^. **Table 2 caption:** **Comparison 1: COVID-19 positive (n=669) vs not COVID-19 positive (tested negative <u>plus</u> untested cohort) (n=501,837); Comparison 2: COVID-19 positive (n=669) vs COVID-19 test negative (n=805); Comparison 3: COVID-19 test negative (n=805) vs untested population (n=501,032). Results are odds ratio, 95% confidence interval, and p-value (from top to bottom) for each exposure. For continuous variables (age, BMI) coefficients refer to the effect of one per unit increase in the exposures, i.e. 1-year increase in age and 1kg/m^2^ increase in BMI, on odds of the outcomes. The remaining exposures are set as binary measures with results showing effect of change from non-disease to disease states, male sex vs female sex, white ethnicity vs non-white ethnicity; smoking history (current/previous) vs never smoked; ACEi/ARB use vs no ACEi/ARB use on odds of the outcome. *indicates p-values <0.05. ACEi: Angiotensin Converting Enzyme inhibitor; ARB: Angiotensin Receptor Blocker; BMI: body mass index; coronavirus 2019: COVID-19; MI: myocardial infarction.

|  | Comparison 1 |  | Comparison 2 |  | Comparison 3 |  |
| --- | --- | --- | --- | --- | --- | --- |
| *Predictors* | Univariate | Multivariate | Univariate | Multivariate | Univariate | Multivariate |
| Male sex | 1.551* | 1.360* | 1.258* | 1.314* | 1.233* | 1.045 |
|  | [1.331, 1.808] | [1.159, 1.597] | [1.024, 1.546] | [1.055, 1.639] | [1.074, 1.417] | [0.904, 1.208] |
|  | 1.88x10<sup>-08</sup> | 1.63x10<sup>-04</sup> | 0.0292 | 0.0151 | 0.0029 | 0.5517 |
| Age (years) | 1.012* | 1.003 | 0.994 | 0.995 | 1.019* | 1.007 |
|  | [1.002, 1.022] | [0.993, 1.014] | [0.982, 1.006] | [0.982, 1.009] | [1.011, 1.028] | [0.997, 1.017] |
|  | 0.0148 | 0.5413 | 0.3114 | 0.4817 | 1.88x10<sup>-05</sup> | 0.1551 |
| White ethnicity | 0.329* | 0.326* | 0.467* | 0.488* | 0.704* | 0.685* |
|  | [0.267, 0.410] | [0.261, 0.411] | [0.331, 0.653] | [0.340, 0.696] | [0.546, 0.926] | [0.525, 0.912] |
|  | 3.48x10<sup>-24</sup> | 3.83x10<sup>-22</sup> | 1.09x10<sup>-05</sup> | 8.75x10<sup>-05</sup> | 0.0090 | 0.0073 |
| BMI (kg/m<sup>2</sup>) | 1.064* | 1.044* | 1.026* | 1.021* | 1.036* | 1.017* |
|  | [1.050, 1.077] | [1.029, 1.060] | [1.008, 1.046] | [1.001, 1.042] | [1.022, 1.049] | [1.003, 1.032] |
|  | 2.58x10<sup>-20</sup> | 1.11x10<sup>-08</sup> | 0.0057 | 0.0404 | 1.50x10<sup>-07</sup> | 0.0203 |
| Diabetes | 2.514* | 1.396* | 1.311 | 1.192 | 1.920* | 1.253 |
|  | [2.050, 3.057] | [1.100, 1.760] | [0.989, 1.740] | [0.849, 1.673] | [1.564, 2.333] | [0.992, 1.571] |
|  | 1.49x10<sup>-19</sup> | 0.0053 | 0.0597 | 0.3100 | 1.61x10<sup>-10</sup> | 0.0544 |
| Hypertension | 1.890* | 1.390* | 1.063 | 1.01 | 1.780* | 1.368* |
|  | [1.624, 2.200] | [1.143, 1.688] | [0.866, 1.305] | [0.771, 1.322] | [1.550, 2.044] | [1.144, 1.633] |
|  | 1.91x10<sup>-16</sup> | 9.17x10<sup>-04</sup> | 0.559 | 0.9443 | 3.13x10<sup>-16</sup> | 5.54x10<sup>-04</sup> |
| High cholesterol | 1.831* | 1.130 | 1.078 | 1.02 | 1.700* | 1.133 |
|  | [1.561, 2.142] | [0.923, 1.379] | [0.870, 1.337] | [0.772, 1.347] | [1.467, 1.964] | [0.941, 1.361] |
|  | 6.58x10<sup>-14</sup> | 0.2326 | 0.4923 | 0.8906 | 1.00x10<sup>-12</sup> | 0.1852 |
| ACEi/ARB use | 1.682* | 0.898 | 1.075 | 0.956 | 1.566* | 0.919 |
|  | [1.395, 2.014] | [0.715, 1.127] | [0.836, 1.380] | [0.695, 1.316] | [1.316, 1.853] | [0.744, 1.134] |
|  | 2.90x10<sup>-08</sup> | 0.3558 | 0.5737 | 0.7841 | 2.69x10<sup>-07</sup> | 0.4337 |
| Prior MI | 2.117* | 1.313 | 0.805 | 0.702 | 2.635* | 1.821* |
|  | [1.592, 2.757] | [0.965, 1.756] | [0.562, 1.147] | [0.470, 1.042] | [2.081, 3.290] | [1.403, 2.335] |
|  | 8.06x10<sup>-08</sup> | 0.0738 | 0.2342 | 0.0812 | 1.05x10<sup>-16</sup> | 3.84x10<sup>-06</sup> |
| Smoking | 1.426* | 1.374* | 0.935 | 0.982 | 1.526* | 1.412* |
|  | [1.223, 1.662] | [1.172, 1.611] | [0.760, 1.150] | [0.788, 1.224] | [1.328, 1.755] | [1.223, 1.631] |
|  | 5.67x10<sup>-06</sup> | 8.78x10<sup>-05</sup> | 0.5227 | 0.8729 | 2.78x10<sup>-09</sup> | 2.71x10<sup>-06</sup> |

**Figure 2.**
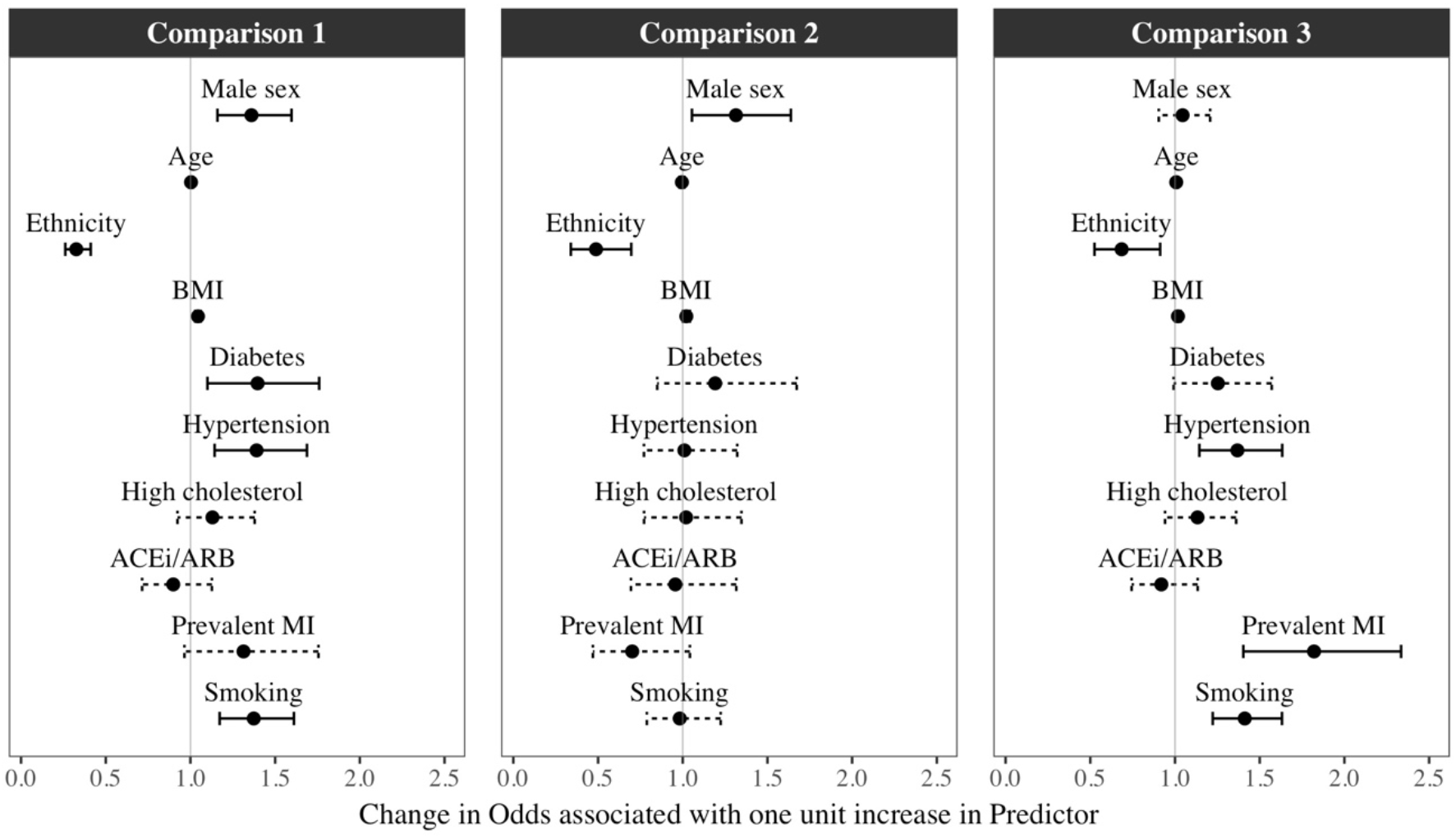
Odds Ratios and 95% confidence intervals for each exposure from the multivariate logistic regression models in the three different comparisons*. **Figure 2 legend:** “Comparison 1: COVID-19 positive (n=669) vs not COVID-19 positive (tested negative <u>plus</u> untested cohort) (n=501,837); Comparison 2: COVID-19 positive (n=669) vs COVID-19 test negative (n=805); Comparison 3: COVID-19 test negative (n=805) vs untested population (n=501,032). Results are odds ratios with 95% confidence intervals. Dashed lines represent non-significant and solid lines statistically significant results, with threshold at p<0.05.

#### COVID-19 positive vs COVID-19 tested negative

We next considered associations between exposures and COVID-19 positives (n=669) vs tested negative cohort (n=805). Within this sample, the univariate predictors of positivity were male sex, nonwhite ethnicity, and greater BMI. These variables remained statistically significant in the multivariate model with adjustment for all other covariates (Table 2, Figure 2). The greatest magnitude of effect related to ethnicity, with white ethnicity associated with 51% lower odds of COVID-19 positive status than other ethnicities [OR 0.488, CI (0.340, 0.696)]. Compared with women, men had 31% greater odds of a COVID-19 positive test [OR 1.314, CI (1.055, 1.639)] and for every 1kg/m^2^ of BMI, there was 2% greater odds of COVID-19 positive status (Table 2, Figure 2).

#### COVID-19 tested negatives vs untested population

Finally, we investigated associations between the exposures and tested negatives (n=805) vs untested UKB population (n=501,032). There were significant univariate associations for all covariates considered. In the multivariate model, non-white ethnicity, higher BMI, hypertension, previous MI, and smoking were significant predictors of a having a negative test, and therefore of presenting to hospital, perhaps with respiratory symptoms, compared to not being tested (Table 2, Figure 2).

## DISCUSSION

### Summary of findings

In this analysis of the first release of 1,474 COVID-19 test results from the UKB, non-white ethnicity, male sex, greater BMI, diabetes, hypertension, prior MI and smoking were independently associated with COVID-19 positive test in comparison to rest of the cohort (tested negatives plus untested). However, within the tested cohort, a positive result was more likely for men, those of non-white ethnicity, and with greater BMI. Indeed, when compared with the background population, the pattern of associations between exposures and COVID-19 positive was similar to that for COVID-19 test negative. These findings suggest that non-white ethnicity, male sex, and higher BMI have specific relevance to COVID-19, whilst the associations between COVID-19 positive and the remainder of the population reflect morbidities associated with general requirement for hospitalisation, without specificity to COVID-19. Furthermore, as testing was almost fully limited to hospitalised patients at this time, these associations relate specifically to the more severe end of the COVID-19 disease manifestations, requiring hospitalisation. Notably, ACEi/ARB usage was not associated with COVID-19 status.

### Comparison with existing literature

With the rapid global spread of COVID-19, understanding the determinants of infection risk and severity is a priority. Differences in ethnic background are known to contribute to differences in patterns of a number of diseases, including influenza^12^, due to different genetic susceptibilities and environmental exposures^13^. In global surveillance data, disparities in COVID-19 mortality rates across different countries are striking, but the interpretation of this data is limited by widely differing testing policies, healthcare systems, resources, and infection control policies. In the UK, national audit data demonstrates as many as one-third of COVID-19 patients admitted to intensive care are from black and minority ethnic backgrounds (BME); a rate which is disproportionate to BME’s representation among the general UK population^14^. In our study, ethnicity appeared to have specific association with COVID-19 positive status that appeared independent from the often-quoted confounders of cardiovascular and metabolic morbidity that are known to be higher in prevalence in BME cohorts^15^. Having accounted for cardiometabolic morbidity, the possible explanations for this association remain numerous^16^, gravitating around both genetic and social factors: behavioural, cultural, and socioeconomic differences, including health-seeking behaviour and intergenerational cohabitation are all likely to play a role in the strong disparity observed in our study, providing key targets for both further research and public health policy.

Since the first reports emerging from China at the beginning of the outbreak, it has been widely recognized that males suffer higher rates of infection and poorer outcomes compared to females; with reported distributions of approximately three-fifths men and two-fifths women^17,18^. The reasons for this are unclear. Animal studies demonstrate, that in mice infected with SARS-CoV, oestrogen-deplete status either due to male gender or ovariectomy is associated with higher risk of acute respiratory distress syndrome (ARDS), indicating a possible protective role of oestrogen signalling^19^. Men are known to have higher burden of cardiovascular disease than women up to the perimenopausal years; and thus, lower cardiometabolic morbidity among women in the younger cohort has been postulated to contribute to better outcomes. However, we demonstrate that in our study population, the association between male sex and higher infection rates was independent of cardiometabolic disease. Furthermore, male sex appears significant in our sample comprising an older cohort with almost all women being post-menopause, indicating that sex-differential disparities in COVID-19 disease severity relate to factors other than immediate-term oestrogen exposure. Thus, our findings suggest that the higher risk of COVID-19 in men is not sufficiently explained by the oestrogen pathway or greater burden of cardiometabolic disease.

Obesity is a global health issue, rising in prevalence and public health burden in both developed and developing countries. Patients who suffer from obesity are known to be at increased risk of a number of conditions, including cardiometabolic and respiratory disease, contributing to a poor physiological reserve. It is already known that patients with obesity have worse outcomes from influenza infection^20,21^. With the wealth of emerging research on COVID-19, concern has grown over the association between obesity and poor outcomes of infection^22^; with studies consistently demonstrating higher rates of critical or intensive care requirement among individuals with higher BMI^23–25^. Similar to ethnicity, the relationship between obesity and severe infection must be isolated from the confounding of obesity-related comorbidity. In our study, we demonstrate the distinct role of obesity from that of associated cardiometabolic diseases; with the major finding that obesity, and not its comorbidities, had independent and specific association with COVID-19 positivity. This is of important relevance, as mechanistic understanding of the reason behind this association may provide therapeutic insight. For example, obesity enhances risk of thrombosis, which has been a recent focus of interest given concern over a possible association between COVID-19 and prothrombotic intravascular coagulation^26^. The results of our study provide useful information for risk stratification of patients, highlight important avenues for further research, and emphasise the public health-level importance of continued targeting of obesity.

Several reports hypothesise potential mechanistic links between ACEi/ARB usage and adverse outcomes from COVID-19^5^. SARS-CoV-2 has been shown to exhibit specific tropism for the angiotensin-converting enzyme 2 (ACE2) receptor; by which means it enters the cells and establishes itself in the host^27^. The expression of ACE2 receptors in epithelial cells of the lung, intestine, kidney and endothelium may be increased in those treated with ACEi/ARBs, thereby facilitating entry and multisystem manifestations of COVID-19^28,29^. The relationship between COVID19 infection risk and use of ACEi/ARBs has been a matter of debate since the early days of the outbreak, but recent studies have revealed a lack of independent association when morbidity variables, including atherosclerotic cardiovascular disease, heart failure and cardiometabolic diseases such as diabetes and hypertension were accounted for^5,6^. Findings from our sample are consistent with these later reports, demonstrating a univariate association with ACEi/ARB use which becomes non-significant after adjustment for cardiometabolic and demographic factors.

### Strengths and Limitations

The UK Biobank is a comprehensive data source, incorporating a large sample with linkages to prospectively tracked health outcomes recorded in a standardised manner using ICD codes, enabling reliable and up-to-date definition of morbidities. The rapid release of COVID-19 testing data provides a huge opportunity to examine association of a large number of exposures with COVID-19 status and outcomes. As this was the very first release of this data, the COVID-19 positive caseload was low; this dataset is continuously updated and there may be opportunity in the future to consider a wider range of exposures in larger samples. The low number of COVID-19 cases reflects both the UK national testing policy, in accordance with which individuals not requiring hospital admission were not tested, and the novelty of the disease that precludes any sources of long-term data collection. Due to the observational study design, we cannot comment on causal relationships from the results, however, the prospective nature of the study ensures confident temporal separation of exposure and outcome. Finally, the aforementioned testing policy in act during the time of data collection presents a further limitation to the study. Individuals who were infected with COVID-19 will only have been tested if they were unwell enough to present to hospital; the ‘positive’ cohort thus mostly represents cases of moderate or high severity. Therefore, whilst this selective approach to testing allow inferences regarding associations with more severe manifestations of the disease, the results of the study may not be generalisable to asymptomatic, or only mildly symptomatic patients.

### Conclusions

This work highlights specific associations of non-white ethnicity, male sex, and higher body mass index with COVID-19 positive status, which were independent of other demographic or cardiometabolic factors. More detailed characterisation of these associations in larger and more diverse cohorts is warranted, particularly with regards ethnicity. Investigation of potential biological pathways underlying these observed associations may provide insight into the mechanisms by which SARS-CoV-2 causes disease enabling more informed pursuit of potential therapeutic targets.

## Data Availability

This analysis has been performed using UK Biobank data under access application 2964. The UK Biobank is an open access research resource with data available to all bone fide researchers on formal request without preferential access or exclusivity.

## References

1. World Health Organization (WHO). Novel Coronavirus (2019-nCoV), situation report-1, 21 January 2020. https://www.who.int/docs/default-source/coronaviruse/situation-reports/20200121-sitrep-1-2019-ncov.pdf (Accessed 8th May 2020)

2. Ruan Q, Yang K, Wang W, Jiang L, Song J. Clinical predictors of mortality due to COVID-19 based on an analysis of data of 150 patients from Wuhan, China. Intensive Care Med. 2020. doi: 10.1007/s00134-020-05991-x

3. Shi S, Qin M, Shen B, Cai Y, Liu T, Yang F, et al. Association of Cardiac Injury with Mortality in Hospitalized Patients with COVID-19 in Wuhan, China. JAMA Cardiol 2020; doi: 10.1001/jamacardio.2020.0950

4. Guo T, Fan Y, Chen M, Wu X, Zhang L, He T, et al. Cardiovascular Implications of Fatal Outcomes of Patients with Coronavirus Disease 2019 (COVID-19). JAMA Cardiol 2020; doi: 10.1001/jamacardio.2020.1017

5. Vaduganathan M, Vardeny O, Michel T, McMurray JJV, Pfeffer MA, Solomon SD. Renin–Angiotensin–Aldosterone System Inhibitors in Patients with Covid-19. N Engl J Med 2020;382:1653–1659. doi: 10.1056/nejmsr2005760

6. Mehra M, Desai S, Kuy S, Henry T, Patel A. Cardiovascular Disease, Drug Therapy, and Mortality in Covid-19. N Engl J Med 2020: in press. doi: 10.1056/NEJMoa2007621

7. Li J, Wang X, Chen J, Zhang H, Deng A. Association of Renin-Angiotensin System Inhibitors With Severity or Risk of Death in Patients With Hypertension Hospitalized for Coronavirus Disease 2019 (COVID-19) Infection in Wuhan, China. JAMA Cardiol 2020;2019:1–6. doi: 10.1001/jamacardio.2020.1624

8. UK Biobank: Protocol for a large-scale prospective epidemiological resource. 2007. https://www.ukbiobank.ac.uk/wp-content/uploads/2011/11/UK-Biobank-Protocol.pdf (Accessed 8th May 2020)

9. Schnier C, Bush K, Nolan J, Sudlow C. Definitions of Acute Myocardial Infarction and Main Myocardial Infarction Pathological Types UK Biobank Phase 1 Outcomes Adjudication on behalf of UK Biobank Outcome Adjudication Group Definitions of Acute Myocardial Infarction. 2017; http://biobank.ndph.ox.ac.uk/showcase/showcase/docs/alg_outcome_mi.pdf (Accessed 8th May 2020)

10. Huang C, Wang Y, Li X, Ren L, Zhao J, Hu Y, et al. Clinical features of patients infected with 2019 novel coronavirus in Wuhan, China. Lancet 2020;395:497–506. doi: 10.1016/S0140-6736(20)30183-5

11. Wang D, Hu B, Hu C, Zhu F, Liu X, Zhang J, et al. Clinical Characteristics of 138 Hospitalized Patients with 2019 Novel Coronavirus-Infected Pneumonia in Wuhan, China. JAMA - J Am Med Assoc 2020;323:1061–1069. doi: 10.1001/jama.2020.1585

12. Zhao H, Harris RJ, Ellis J, Pebody RG. Ethnicity, deprivation and mortality due to 2009 pandemic influenza A(H1N1) in England during the 2009/2010 pandemic and the first post-pandemic season. Epidemiol Infect 2015;143:3375–3383. doi: 10.1017/S0950268815000576

13. Lee C. ‘Race’ and ‘ethnicity’ in biomedical research: How do scientists construct and explain differences in health? Soc Sci Med 2009;68:1183–1190. doi: 10.1016/j.socscimed.2008.12.036

14. ICNARC report on COVID-19 in critical care. ICNARC COVID-19 Study Case Mix Program Database. 2020. https://www.icnarc.org/Our-Audit/Audits/Cmp/Reports (Accessed 8th May 2020)

15. Tillin T, Hughes AD, Mayet J, Whincup P, Sattar N, Forouhi NG, et al. The relationship between metabolic risk factors and incident cardiovascular disease in Europeans, South Asians, and African Caribbeans: SABRE (Southall and Brent Revisited) - A prospective population-based study. J Am Coll Cardiol 2013;61:1777–1786. doi: 10.1016/j.jacc.2012.12.046

16. Pareek M, Bangash MN, Pareek N, Pan D, Sze S, Minhas JS, et al. Ethnicity and COVID-19: an urgent public health research priority. Lancet. 2020;395:1421–1422.

17. Richardson S, Hirsch JS, Narasimhan M, Crawford JM, McGinn T, Davidson KW, et al. Presenting Characteristics, Comorbidities, and Outcomes Among 5700 Patients Hospitalized With COVID-19 in the New York City Area. JAMA American Medical Association (AMA); Published online 22^nd^ April 2020; doi: 10.1001/jama.2020.6775

18. Guan W, Ni Z, Hu Y, Liang W, Ou C, He J, et al. Clinical Characteristics of Coronavirus Disease 2019 in China. N Engl J Med 2020;382:1708–1720. doi: 10.1056/NEJMoa2002032

19. Channappanavar R, Fett C, Mack M, Eyck PP Ten, Meyerholz DK, Perlman S. Sex-Based Differences in Susceptibility to Severe Acute Respiratory Syndrome Coronavirus Infection. J Immunol 2017;198:4046–4053. doi: 10.4049/jimmunol.1601896

20. Green WD, Beck MA. Obesity Impairs the Adaptive Immune Response to Influenza Virus. Ann Am Thorac Soc 2017;14:406–409. doi: 10.1513/AnnalsATS.201706-447AW

21. Luzi L, Radaelli MG. Influenza and obesity: its odd relationship and the lessons for COVID-19 pandemic. Acta Diabetol 2020; doi: 10.1007/s00592-020-01522-8

22. Sattar N, McInnes IB, McMurray JJV. Obesity a Risk Factor for Severe COVID-19 Infection: Multiple Potential Mechanisms. Circulation 2020; doi: 10.1161/circulationaha.120.047659

23. Petrilli CM, Jones SA, Yang J, Rajagopalan H, O’Donnell LF, Chernyak Y, et al. Factors associated with hospitalization and critical illness among 4,103 patients with COVID-19 disease in New York City. *medRxiv* 2020; doi: 2020.04.08.20057794.

24. Simonnet A, Chetboun M, Poissy J, Raverdy V, Noulette J, Duhamel A, Labreuche J, Mathieu D, Pattou F, Jourdain M. High prevalence of obesity in severe acute respiratory syndrome coronavirus-2 (SARS-CoV-2) requiring invasive mechanical ventilation. Obesity 2020; in press. doi: 10.1002/oby.22831

25. Lighter J, Phillips M, Hochman S, Sterling S, Johnson D, Francois F, Stachel A. Obesity in patients younger than 60 years is a risk factor for Covid-19 hospital admission. Clin Infect Dis; ciaa415. doi: 10.1093/cid/ciaa415

26. Tang N, Li D, Wang X, Sun Z. Abnormal coagulation parameters are associated with poor prognosis in patients with novel coronavirus pneumonia. J Thromb Haemost 2020;18:844-847. doi: 10.1111/jth.14768

27. Hoffmann M, Kleine-Weber H, Schroeder S, Krüger N, Herrler T, Erichsen S, et al. SARS-CoV-2 Cell Entry Depends on ACE2 and TMPRSS2 and Is Blocked by a Clinically Proven Protease Inhibitor. Cell 2020;181:271–280 doi: 10.1016/j.cell.2020.02.052

28. Wan Y, Shang J, Graham R, Baric RS, Li F. Receptor Recognition by the Novel Coronavirus from Wuhan: an Analysis Based on Decade-Long Structural Studies of SARS Coronavirus. J Virol 2020;94:e00127–20. doi: 10.1128/jvi.00127-20

29. Li XC, Zhang J, Zhuo JL. The vasoprotective axes of the renin-angiotensin system: Physiological relevance and therapeutic implications in cardiovascular, hypertensive and kidney diseases. Pharmacol Res 2017;125:21–38. doi: 10.1016/j.phrs.2017.06.005

30. Li B, Yang J, Zhao F, Zhi L, Wang X, Liu L, et al. Prevalence and impact of cardiovascular metabolic diseases on COVID-19 in China. Clin Res Cardiol 2020;109:531–538. doi: 10.1007/s00392-020-01626-9

31. Yang J, Zheng Y, Gou X, Pu K, Chen Z, Guo Q, et al. Prevalence of comorbidities and its effects in coronavirus disease 2019 patients: A systematic review and meta-analysis. Int J Infect Dis 2020;94:91–95.

